# The decrease in hospitalizations for transient ischemic attack and ischemic stroke, especially in mild cases, during the COVID-19 epidemic in Japan

**DOI:** 10.1101/2020.11.17.20233692

**Authors:** Hiroyuki Nagano, Daisuke Takada, Jung-ho Shin, Tetsuji Morishita, Susumu Kunisawa, Yuichi Imanaka

**Affiliations:** Department of Healthcare Economics and Quality Management, Graduate School of Medicine, Kyoto University, Kyoto City, Japan

## Abstract

**Background and Purpose:** The epidemic of the coronavirus disease 2019 (COVID-19) has affected health care systems globally. The aim of our study was to assess the impact of the COVID-19 epidemic on hospital admissions for stroke in Japan.

**Methods:** We analyzed administrative (Diagnosis Procedure Combination) data for cases of inpatients aged 18 years and older who were diagnosed with stroke (ischemic stroke, transient ischemic attack (TIA), hemorrhagic stroke, or subarachnoid hemorrhage (SAH)) and discharged from hospital during the period July 1, 2018 to June 30, 2020. The number of patients with each stroke diagnosis, various patient characteristics, and treatment approaches were compared before and after the epidemic. Changes in the trend of the monthly number of inpatients with each stroke diagnosis were assessed using interrupted time-series analyses.

**Results:** A total of 111,922 cases (ischemic stroke: 74,897 cases; TIA: 5,374 cases; hemorrhagic stroke: 24,779 cases; SAH: 6,872 cases) in 253 hospitals were included. The number of cases for all types of stroke decreased (ischemic stroke: -13.9%; TIA: -21.4%; hemorrhagic stroke: -9.9%; SAH: -15.2%) in April and May 2020, compared to the number of cases in 2019. Ischemic stroke and TIA cases, especially mild cases (modified Rankin Scale = 0), decreased, with a statistically significant change in trend between the before- and after-epidemic periods.

**Conclusions:** These data showed a marked reduction in the number of hospital admissions due to stroke during the COVID-19 epidemic. The change in Ischemic stroke and TIA cases, especially mild cases, was statistically significant.

## Introduction

The coronavirus disease 2019 (COVID-19), first recognized in Wuhan, China, in early December 2019[1], spread globally in 2020. In Japan, the daily number of new COVID-19 infections increased dramatically from late March and April 2020[2]. In response, the Japanese government declared a state of emergency on April 7 for seven prefectures and broadened the declaration to all 47 prefectures on April 16[3].

Stroke is the second leading cause of death and a major cause of disability in the world[4]. Treatments for stroke, especially intravenous thrombolysis and endovascular intervention for ischemic stroke, are time-sensitive and should be initiated as quickly as possible[5]. Therefore, patients with symptoms suggesting stroke should be evaluated by medical staff immediately. However, movement restrictions and the fear of COVID-19 transmission in hospitals during the COVID-19 epidemic may lead to a hesitation by individuals to seek medical services, resulting in delayed access and a higher risk of severe illness[6-7]. Although previous studies have shown a decline in hospital admissions for stroke during the COVID-19 epidemic[8–11], most of these studies were conducted in a single center or in relatively few centers. In contrast, we performed a retrospective cohort study using a large-scale Japanese database. Our study sought to provide a comprehensive evaluation of the impact of the COVID-19 epidemic on hospital admissions for stroke in Japan.

## Methods

### Data source

For this retrospective cohort study, we extracted Diagnosis Procedure Combination (DPC) data from the Quality Indicator/Improvement Project (QIP) database for inpatients with a date of discharge between July 1, 2018 and June 30, 2020. The QIP database is administered by the Department of Healthcare Economics and Quality Management, Kyoto University[12], which regularly collects DPC data from acute care hospitals in Japan which voluntarily participate in the project. Over 500 hospitals participate in QIP, covering virtually all of Japan. These hospitals include both public and private facilities of various sizes. In 2019, the number of beds at the participating hospitals ranged from 30 to 1,151 (excluding psychiatric, infectious diseases, and tuberculosis beds according to the Japanese classification of hospital beds). The DPC/per-diem payment system (PDPS) is a Japanese prospective payment system applied to acute care hospitals. In 2018, a total of 1,730 hospitals were using the DPC/PDPS, accounting for 54% (482,618 of 891,872) of all the general hospital beds in Japanese hospitals[13-14]. The DPC data consist of claims and discharge summaries, including International Classification of Diseases and Related Health problems 10^th^ Revision (ICD-10) codes classifying the main diagnosis, the cause of hospital admission, the most and second-most medical-resource-intensive diagnoses, up to 10 comorbidities, and 10 complications during hospitalization[15]. The database also includes the following patient details: age, sex, medical procedures, daily records of drug administration, modified Rankin Scale (mRS) before hospital admission and at discharge.

### Study population

We selected cases in which patients aged 18 years and older were hospitalized due to stroke and were discharged between July 1, 2018, and June 30, 2020. Ischemic stroke, transient ischemic attack (TIA), hemorrhagic stroke, and subarachnoid hemorrhage (SAH) were identified by ICD-10 code I63.x, G45.x other than G454, I60.x and I61.x, respectively, as the diagnoses that caused the hospital admission.

### Statistical Analysis

We began with a descriptive analysis. In order to make year-on-year comparisons, we divided the study population into two groups: those who were admitted and discharged from July 2018 to June 2019 and those who were admitted from July 2019 to June 2020. Because the DPC data were coded on the day of discharge, the data for patients who were not discharged by the end of June 2020 were not available, even though their dates of hospital admissions were before June 30, 2020. Thus, both one-year datasets included inpatient cases that started and ended during the corresponding period. We assessed the monthly ratio of cases with each type of stroke between the two groups. In addition, we compared inpatient cases for each diagnosis, patient characteristics, and time-sensitive treatment approaches (intravenous thrombolysis and endovascular intervention) for the months of April and May in 2019 and 2020. We chose this period to compare the effect of the COVID-19 epidemic because the state of emergency was lifted on May 25, 2020, in response to the decrease of new confirmed COVID-19 cases. Continuous variables (such as age) are expressed as median values [interquartile range]. Comparisons of the continuous variables for the two groups were performed using the Mann-Whitney U test. Categorical data (such as sex) were compared using the chi-square or Fisher’s exact test, as appropriate.

An interrupted time series analysis (ITS) was performed to evaluate the effect of the COVID-19 epidemic on the monthly rate of stroke hospitalizations[16]. We statistically tested the changes in the number of admissions based on the date of discharge, after adjusting for seasonality using a Fourier term. We compared the number of admissions per month based on the date of discharge, not the date of admission, in our ITS analyses because, as previously noted, the DPC data were generated after the patient’s discharge from the hospitals. We hypothesized that COVID-19 would impact the admissions volume immediately after April 2020, since the Japanese government had declared the state of emergency and requested self-quarantine and social distancing on April 7, 2020, in response to the increasing number of infected cases. All statistical analyses were performed with R version 3.6.0 (R Foundation for Statistical Computing, Vienna, Austria).

## Results

A total of 111,922 cases (ischemic stroke: 74,897 cases; TIA: 5,374 cases; hemorrhagic stroke: 24,779 cases; SAH: 6,872 cases) in 253 hospitals were included. Among these, 102,685 cases (51,453 and 51,232 cases for the previous and the second years, respectively) were used for year-over-year comparisons (Figure 1). For all types of stroke, the number of cases decreased in April 2020 compared with April 2019 (Figure 2). Monthly numbers of cases and year-over-year ratios for each type of stroke are displayed in Supplemental Table 1. The total number of inpatient cases with each stroke diagnosis, characteristics, and time-sensitive treatment approaches in April and May 2019 and 2020 are described in Tables 1 and 2.

**Table 1:**
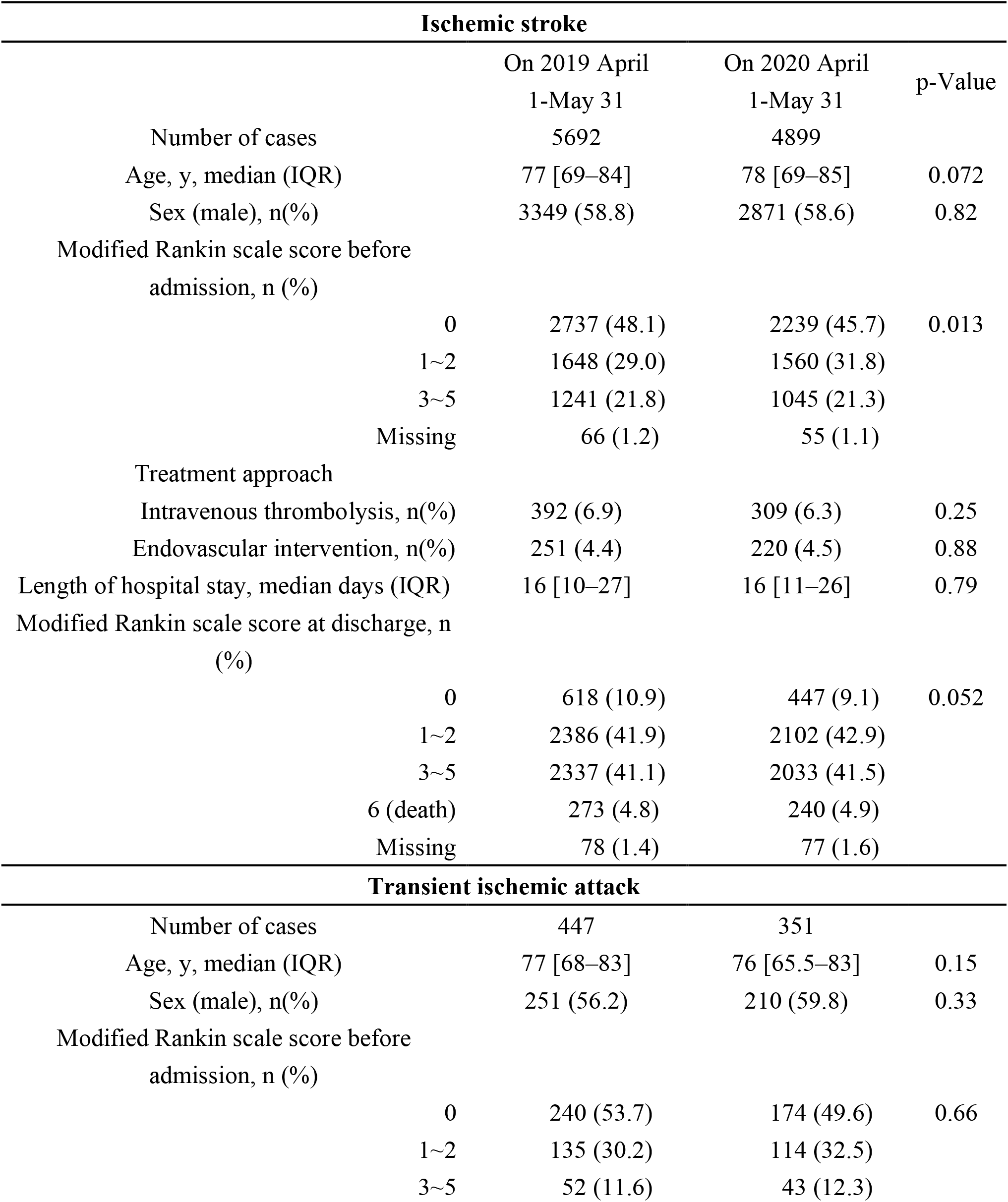
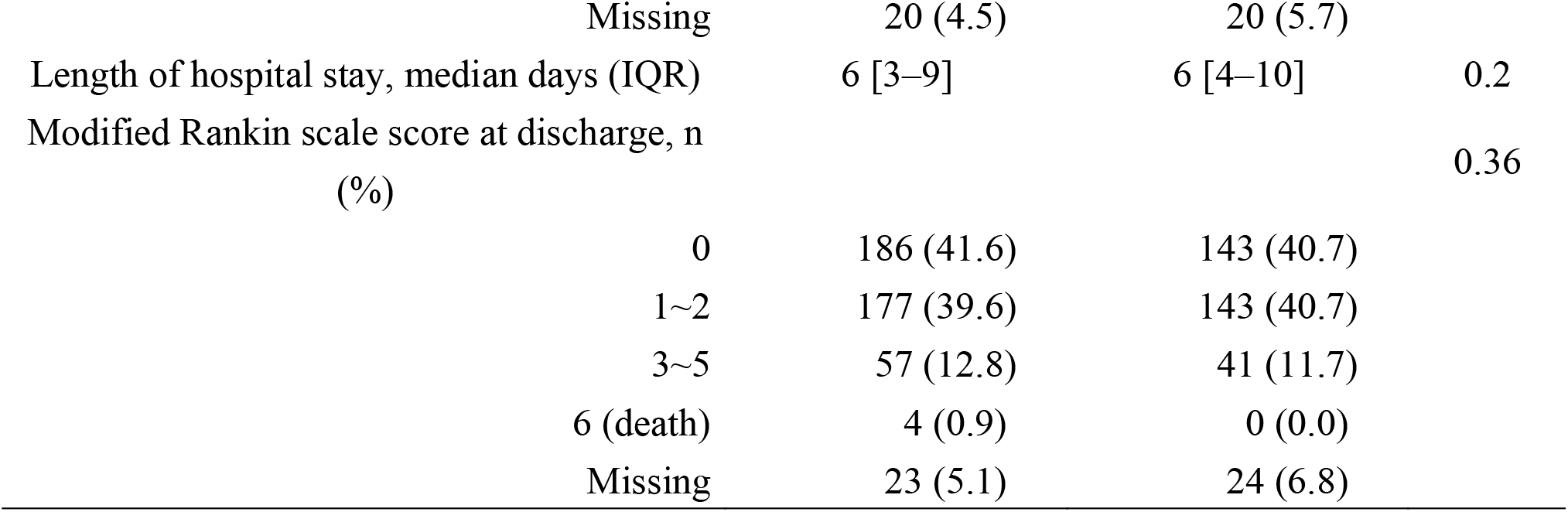
The total number of inpatient cases with ischemic stroke and transient ischemic attack, characteristics, and time-sensitive treatment approaches in April and May of 2019 and 2020.

**Table 2:**
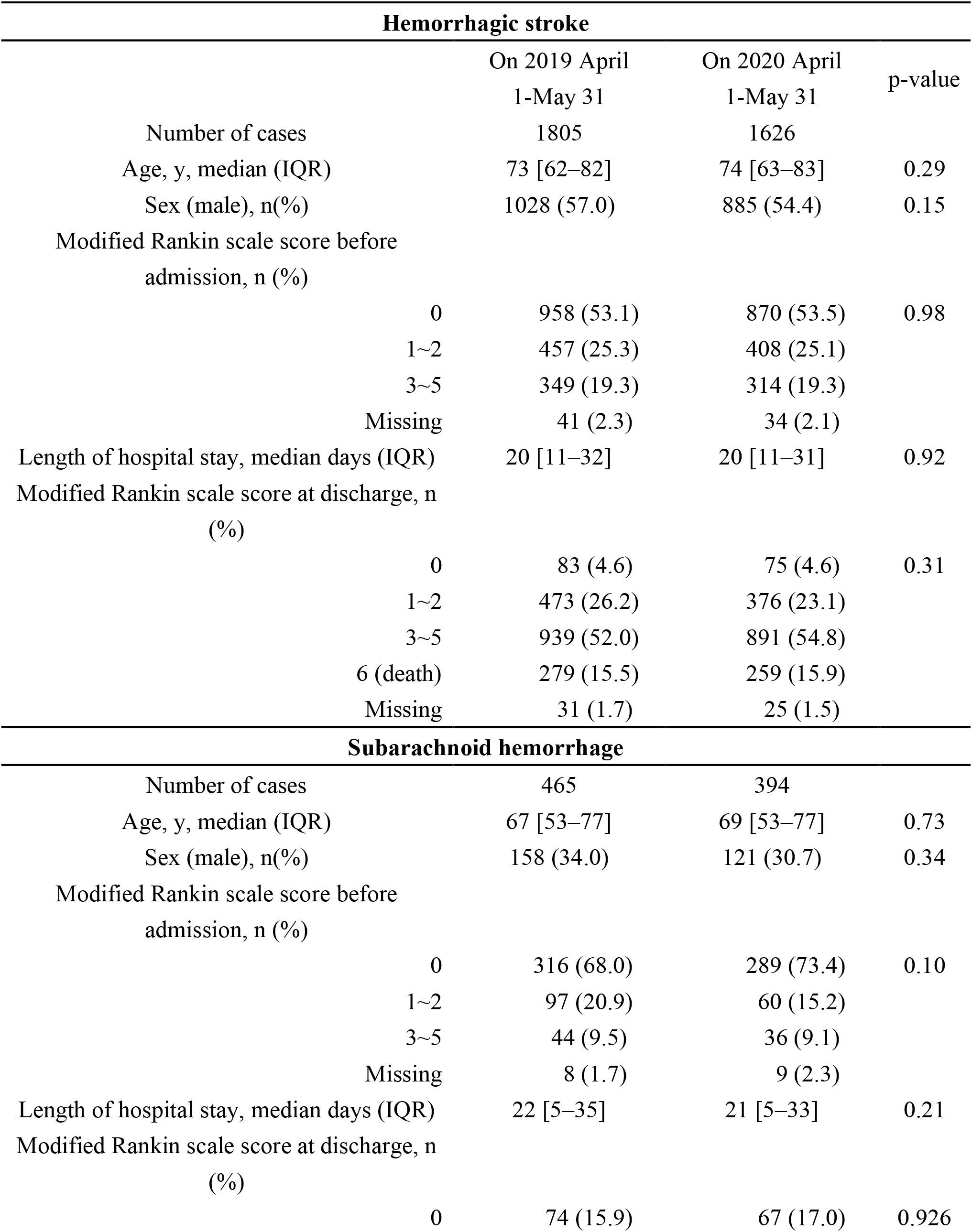
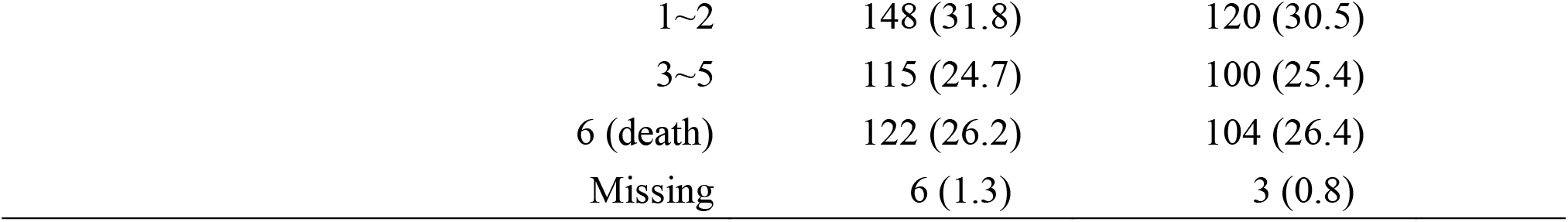
The total number of inpatient cases with hemorrhagic stroke and subarachnoid hemorrhage, and characteristics in April and May of 2019 and 2020.

| Hemorrhagic stroke |  |  |  |  |
| --- | --- | --- | --- | --- |
|  |  | On 2019 April<br>1-May 31 | On 2020 April<br>1-May 31 | p-value |
| Number of cases |  | 1805 | 1626 |  |
| Age, y, median (IQR) |  | 73 [62–82] | 74 [63–83] | 0.29 |
| Sex (male), n(%) |  | 1028 (57.0) | 885 (54.4) | 0.15 |
| Modified Rankin scale score before admission, n (%) |  |  |  |  |
|  | 0 | 958 (53.1) | 870 (53.5) | 0.98 |
|  | 1~2 | 457 (25.3) | 408 (25.1) |  |
|  | 3~5 | 349 (19.3) | 314 (19.3) |  |
|  | Missing | 41 (2.3) | 34 (2.1) |  |
| Length of hospital stay, median days (IQR) |  | 20 [11–32] | 20 [11–31] | 0.92 |
| Modified Rankin scale score at discharge, n (%) |  |  |  |  |
|  | 0 | 83 (4.6) | 75 (4.6) | 0.31 |
|  | 1~2 | 473 (26.2) | 376 (23.1) |  |
|  | 3~5 | 939 (52.0) | 891 (54.8) |  |
|  | 6 (death) | 279 (15.5) | 259 (15.9) |  |
|  | Missing | 31 (1.7) | 25 (1.5) |  |
| Subarachnoid hemorrhage |  |  |  |  |
| Number of cases |  | 465 | 394 |  |
| Age, y, median (IQR) |  | 67 [53–77] | 69 [53–77] | 0.73 |
| Sex (male), n(%) |  | 158 (34.0) | 121 (30.7) | 0.34 |
| Modified Rankin scale score before admission, n (%) |  |  |  |  |
|  | 0 | 316 (68.0) | 289 (73.4) | 0.10 |
|  | 1~2 | 97 (20.9) | 60 (15.2) |  |
|  | 3~5 | 44 (9.5) | 36 (9.1) |  |
|  | Missing | 8 (1.7) | 9 (2.3) |  |
| Length of hospital stay, median days (IQR) |  | 22 [5–35] | 21 [5–33] | 0.21 |
| Modified Rankin scale score at discharge, n (%) |  |  |  |  |
|  | 0 | 74 (15.9) | 67 (17.0) | 0.926 |

|  |  |  |
| --- | --- | --- |
| 1~2 | 148 (31.8) | 120 (30.5) |
| 3~5 | 115 (24.7) | 100 (25.4) |
| 6 (death) | 122 (26.2) | 104 (26.4) |
| Missing | 6 (1.3) | 3 (0.8) |

**Figure 1.**
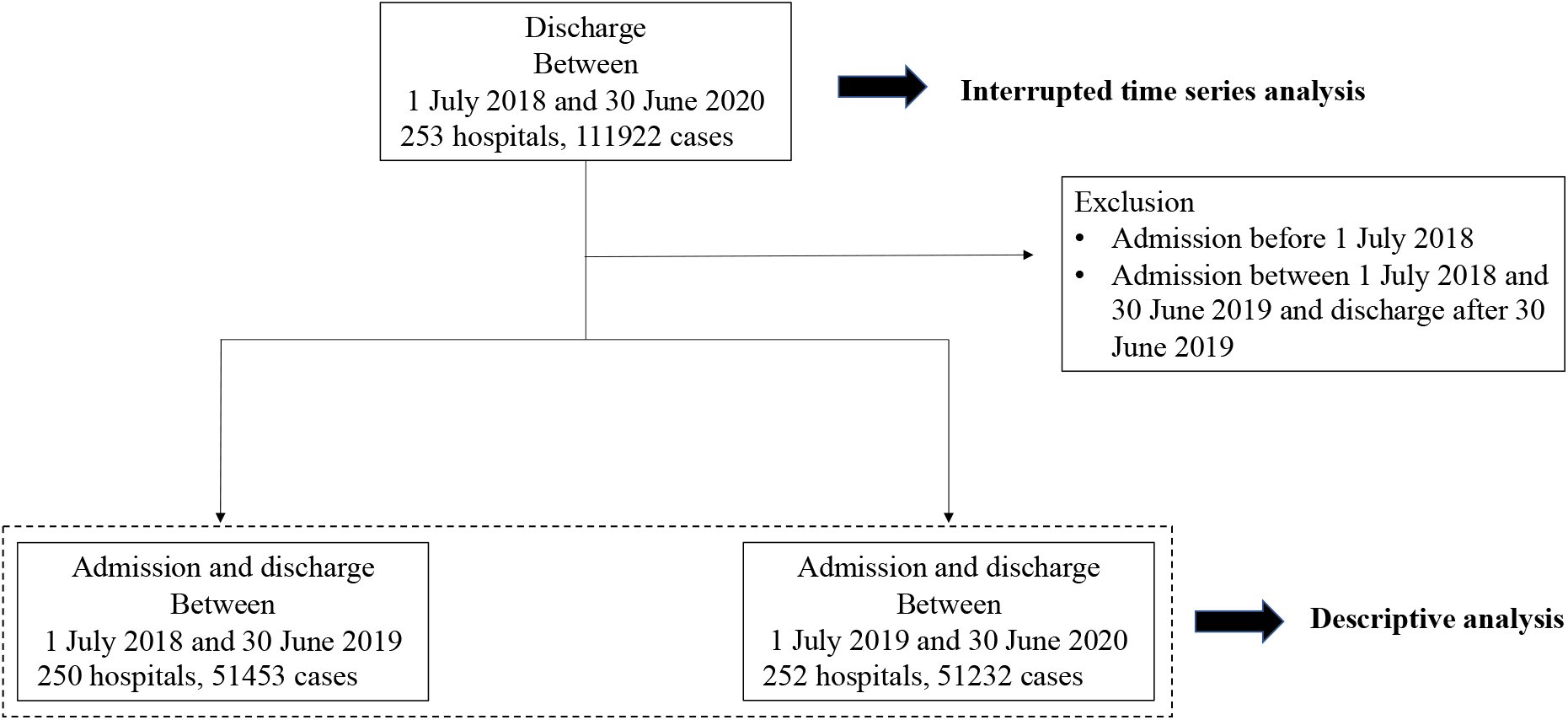
Enrollment and analysis of patients.

**Figure 2.**
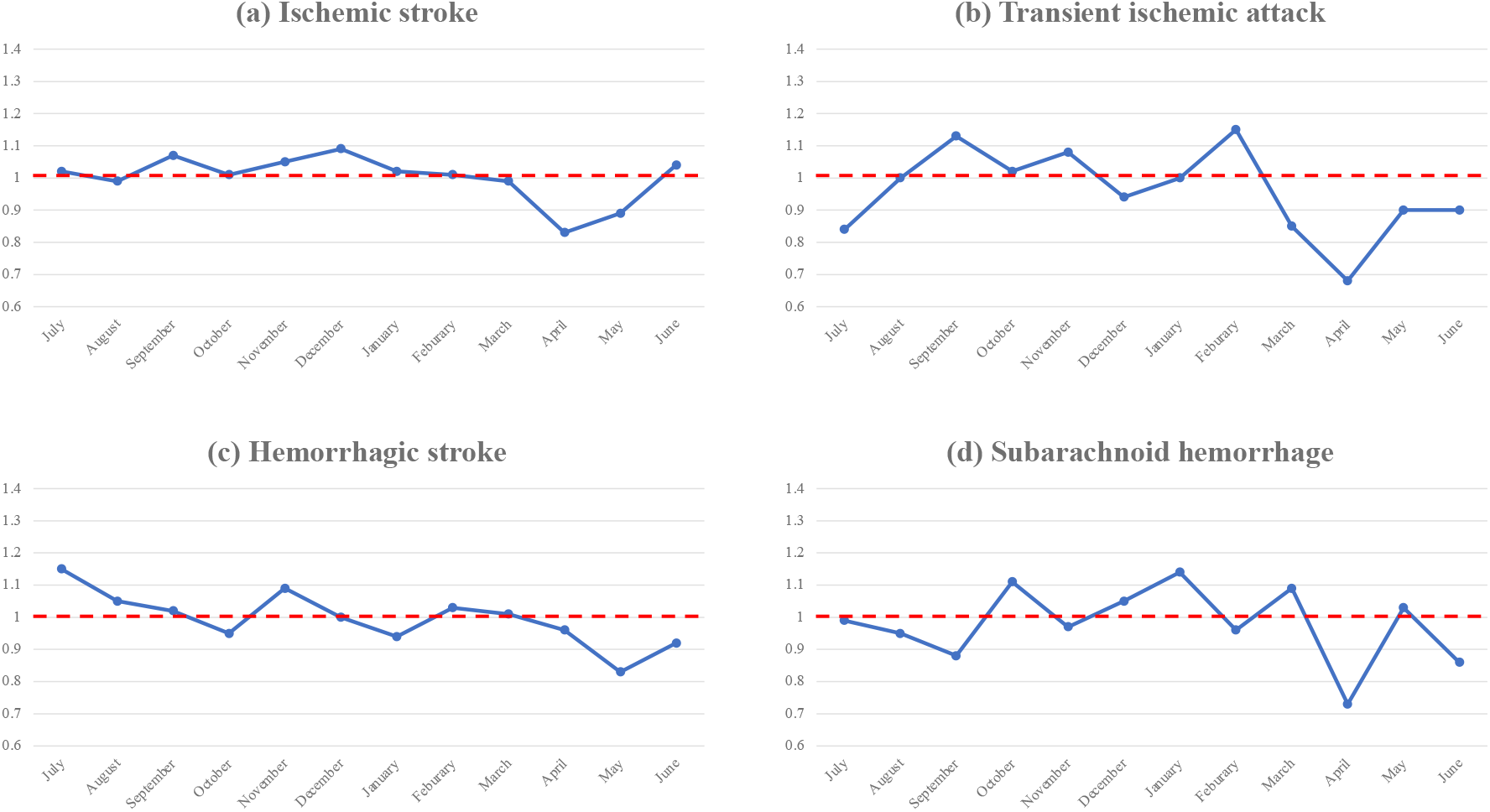
Time trend in year-on-year for each type of stroke. (A) Ischemic stroke; (B) Transient ischemic attack; (C) Hemorrhagic stroke; (D) Subarachnoid hemorrhage. *year-on-year = number of inpatient cases with each diagnosis of stroke per month between July 2019 and June 2020 / number of inpatient cases with each diagnosis of stroke per month between July 2018 and June 2019.

Cases of all types of stroke in April and May 2020 decreased compared with those in 2019 (Ischemic stroke: -13.9%, TIA: -21.4%, Hemorrhagic stroke: -9.9%, SAH: -15.2%). A comparison of the proportion of cases receiving intravenous thrombolysis or endovascular intervention for ischemic stroke remained at the same level (p = 0.2474, p = 0.8773)

The ITS analyses indicated that the COVID-19 epidemic had a negative impact on the number of hospitalizations due to ischemic stroke and TIA (p < 0.05) (Figure 3). The analyses for ischemic stroke and TIA using the mRS indicated that only the number of inpatients with mRS 0 had been negatively impacted by the COVID-19 epidemic. (Table 3)

**Table 3:**
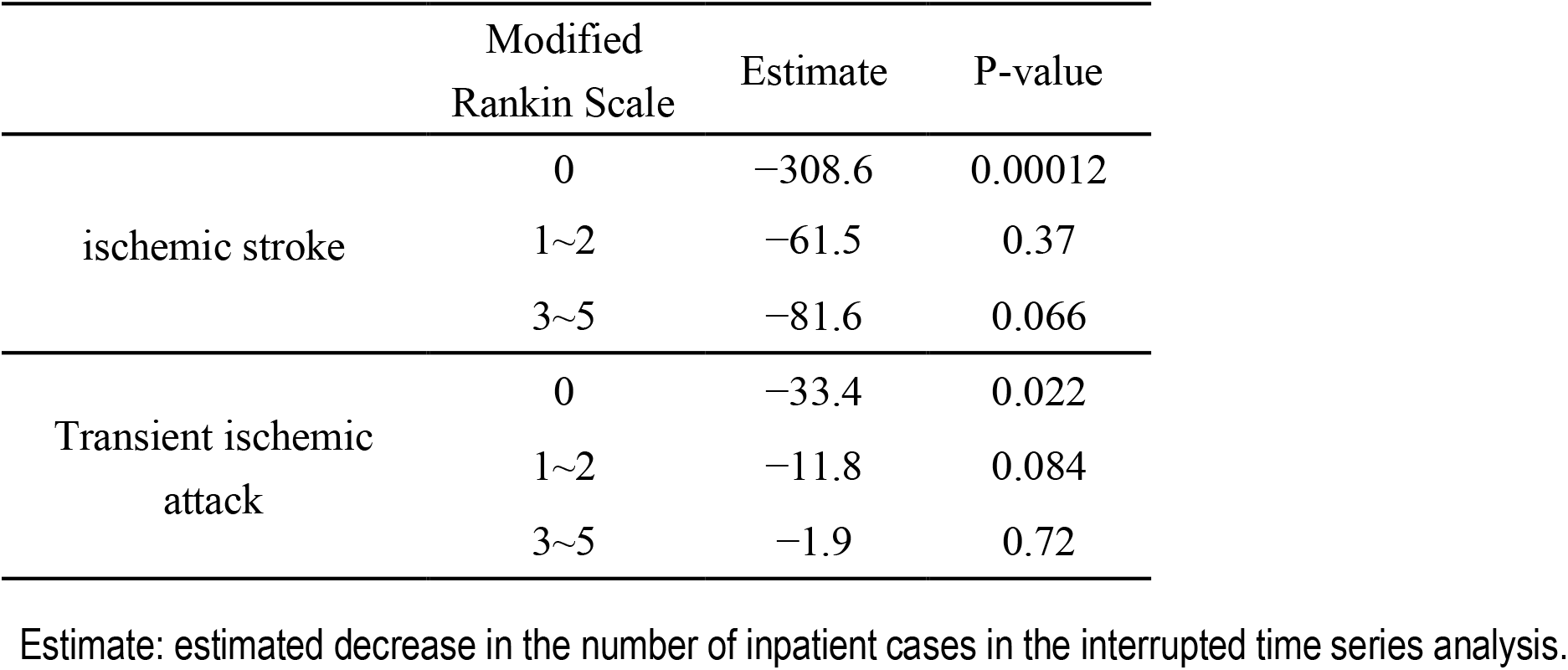
Interrupted time series analysis for admissions cases of ischemic stroke and transient ischemic attack according to the modified Rankin Scale.

|  | Modified Rankin Scale | Estimate | P-value |
| --- | --- | --- | --- |
| ischemic stroke | 0 | −308.6 | 0.00012 |
|  | 1~2 | −61.5 | 0.37 |
|  | 3~5 | −81.6 | 0.066 |
| Transient ischemic attack | 0 | −33.4 | 0.022 |
|  | 1~2 | −11.8 | 0.084 |
|  | 3~5 | −1.9 | 0.72 |

Estimate: estimated decrease in the number of inpatient cases in the interrupted time series analysis.

**Figure 3.**
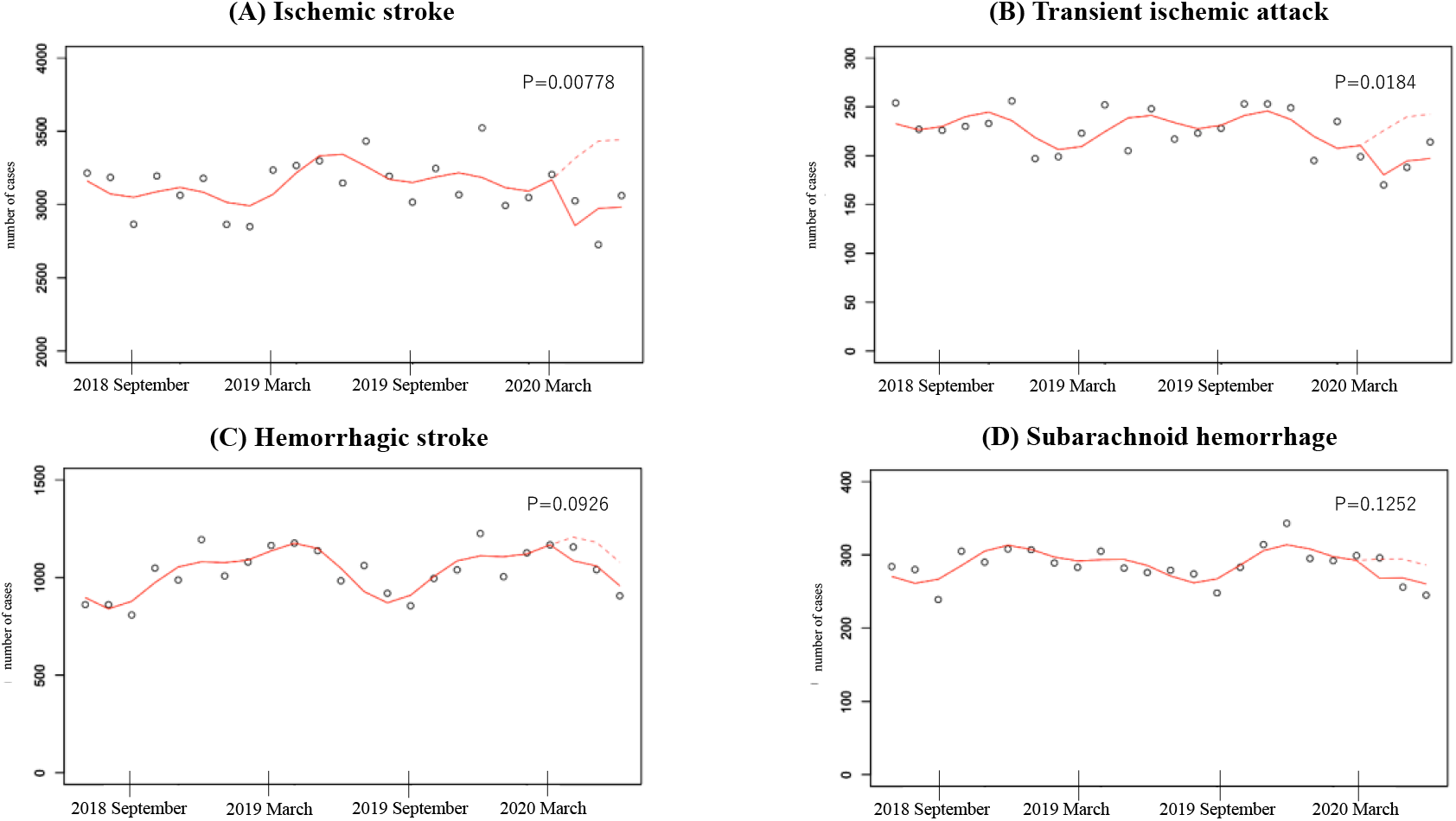
Interrupted time series analysis for the number of inpatients with each diagnosis of stroke between July 2018 and June 2020. Solid lines indicate observed trend following the state of emergency, and dashed lines indicate predicted trend. The p value is shown for each analysis. (A) Ischemic stroke; (B) Transient ischemic attack; (C) Hemorrhagic stroke; (D) Subarachnoid hemorrhage.

## Discussion

In this study, we used a large-scale administrative database to show that the number of inpatients due to all types of stroke decreased during the COVID-19 epidemic in Japan. The ITS analyses identified statistically significant decreases in hospitalizations for ischemic stroke and TIA, especially for mild cases in which the mRS scores were 0.

The significant decrease in the number of inpatient cases due to ischemic stroke and TIA is consistent with previous reports[17]. In other fields, including pediatrics and cardiology, patients were found to be reluctant to seek medical evaluation for fear of exposure to COVID-19 in hospital[7,18]. Similarly, patients of stroke, especially those without severe symptoms such as headaches or paralysis, may be hesitant to seek medical services. Another possible explanation for the decrease in the number of inpatients is the lack of contact with others resulting from stay-at home and social distancing practices. In cases of acute stroke, it is not uncommon for friends and neighbors to request emergency help[19].

Even in emergency conditions such as the COVID-19 epidemic, appropriate evaluation and management of stroke is necessary. Any delay in the medical evaluation of even mild cases may result in severe consequences, including long-term disability, pneumonia due to dysphagia, and the early recurrence of stroke. For example, dual antiplatelet therapy within 24 hours of the onset of symptoms decreases the risk of recurrent stroke and death in patients with high-risk TIA or minor ischemic stroke[20]. In the future, healthcare organizations and providers need to recognize that stroke patients with mild symptoms may be reluctant to seek medical evaluation during an epidemic, particularly where “social distancing” and “stay-at-home” practices are being recommended. Those with symptoms suggesting stroke should be encouraged to seek appropriate medical evaluation immediately, even if the symptoms are mild and temporary.

Our study has several limitations. First, the study population was restricted to cases in hospitals that voluntarily participate in the QIP. The movement of patients to other, non-participating facilities during the COVID-19 epidemic might lead to an overestimate of the impact of the epidemic. However, in the case of hemorrhagic stroke and SAH, the relatively constant trends of hospitalization identified in the study suggest the absence of large-scale movement to other hospitals. It also suggests a lack of significant restrictions on the acceptance of stroke patients by participating hospitals. Second, our DPC data did not include details of the stroke onset time. Consequently, the period of time from symptom onset to hospital arrival was not assessed. Finally, our research included only cases in Japan. Given the differences in healthcare systems and the patterns of the COVID-19 epidemic between countries, it is difficult to judge whether our results can be applied elsewhere. Despite these limitations, our research provides important information on the impact of the COVID-19 epidemic on stroke patients and the facilities caring for them.

## Conclusion

We showed a marked reduction in hospital admissions due to stroke during the COVID-19 epidemic in Japan using large-scale administrative data. In particular, our ITS analyses identified a significant reduction in admissions for ischemic stroke and TIA, especially in mild cases.

## Data Availability

Data are available upon reasonable request. Please contact the corresponding author.

## Contributorship statement

HN: Conceptualization, methodology, software, formal analysis, writing - original draft, writing - review & editing, visualization; DT: Conceptualization, methodology, software, validation, investigation, writing - review & editing; JS: Conceptualization, software, validation, investigation, data curation, writing - review & editing; TM: Conceptualization, methodology, software, validation, investigation, writing - review & editing; SK: Conceptualization, validation, investigation, data curation, resources, writing - review & editing; YI: Conceptualization, validation, investigation, resources, writing - review & editing, supervision, project administration, funding acquisition

## Acknowledgements

We thanked the staff of participating hospitals in QIP.

