## Supplementary material for "The decrease in hospitalizations for transient ischemic attack and ischemic stroke, especially in mild cases, during the COVID-19 epidemic in Japan": in Supplemental Table 1.

SUPPLEMENTAL MATERIAL

table1

| **Ischemic stroke** | July | August | September | October | November | December | January | Feburary | March | April | May | June | Total |
|---|---|---|---|---|---|---|---|---|---|---|---|---|---|
| 2018/7-2019/6 | 3222 | 3154 | 2897 | 3116 | 2977 | 3071 | 3150 | 2882 | 3037 | 2998 | 2694 | 1192 | 34390 |
| 2019/7-2020/6 | 3280 | 3107 | 3109 | 3149 | 3111 | 3338 | 3210 | 2898 | 2992 | 2499 | 2400 | 1241 | 34334 |
| year-on-year | 1.02 | 0.99 | 1.07 | 1.01 | 1.05 | 1.09 | 1.02 | 1.01 | 0.99 | 0.83 | 0.89 | 1.04 | 1.00 |
| **Transient ischemic attack** | | | | | | | | | | | | | |
| 2018/7-2019/6 | 248 | 222 | 229 | 245 | 222 | 251 | 201 | 201 | 233 | 230 | 217 | 191 | 2690 |
| 2019/7-2020/6 | 208 | 221 | 259 | 250 | 240 | 237 | 201 | 232 | 197 | 156 | 195 | 172 | 2568 |
| year-on-year | 0.84 | 1.00 | 1.13 | 1.02 | 1.08 | 0.94 | 1.00 | 1.15 | 0.85 | 0.68 | 0.90 | 0.90 | 0.95 |
| **Hemorrhagic stroke** | | | | | | | | | | | | | |
| 2018/7-2019/6 | 819 | 824 | 882 | 1106 | 1093 | 1154 | 1148 | 1048 | 1057 | 997 | 808 | 316 | 11252 |
| 2019/7-2020/6 | 942 | 868 | 904 | 1050 | 1192 | 1154 | 1083 | 1076 | 1070 | 953 | 673 | 290 | 11255 |
| year-on-year | 1.15 | 1.05 | 1.02 | 0.95 | 1.09 | 1.00 | 0.94 | 1.03 | 1.01 | 0.96 | 0.83 | 0.92 | 1.00 |
| **Subarachnoid hemorrhage** | | | | | | | | | | | | | |
| 2018/7-2019/6 | 272 | 259 | 289 | 297 | 325 | 311 | 284 | 251 | 271 | 278 | 187 | 97 | 3121 |
| 2019/7-2020/6 | 268 | 245 | 254 | 330 | 316 | 326 | 323 | 240 | 296 | 202 | 192 | 83 | 3075 |
| year-on-year | 0.99 | 0.95 | 0.88 | 1.11 | 0.97 | 1.05 | 1.14 | 0.96 | 1.09 | 0.73 | 1.03 | 0.86 | 0.99 |

**The number of inpatient cases per month for each stroke diagnosis.** year-on-year= number of inpatinet cases with each diagnosis of stroke per month between July 2019 and June 2020 / number of inpatient cases with each diagnosis of stroke per month between July 2018 and June 2019
